# Assessing performance and costs of female genital schistosomiasis screening methods in Ondo and Kebbi States, Nigeria

**DOI:** 10.64898/2026.07.12.26357899

**Authors:** Omosefe Osinoiki, Guillaume Trotignon, Akinola S. Oluwole, Martins Imhansoloeva, Anita Jeyam, Iain Jones, Richard Selby, Elena Schmidt

**Affiliations:** Sightsavers Nigeria Country Office, 24 Tennessee Street, off Panama Street, Maitama. FCT, Abuja, Nigeria; Sightsavers, 35 Perrymount Road, Haywards Heath, West Sussex RH16 3BZ, UK

**Keywords:** female genital schistosomiasis, neglected tropical diseases, FGS screening, performance, cost-effectiveness, health system strengthening

## Abstract

Female Genital Schistosomiasis (FGS) is a gynaecological condition, arising from complication from schistosomiasis - a neglected tropical disease (NTD). FGS shares overlapping symptoms with several sexually transmitted infections, making it challenging to diagnose and manage especially in primary health care settings where diagnostic tools are often unavailable. In response to this challenge, an FGS screening tool (COUNTDOWN screening tool) was developed to support primary health care workers in identifying persons at risk of FGS. We investigated the sensitivity and specificity of adapted versions of the COUNTDOWN screening tool and evaluated the costs associated with providing these services in two Nigerian schistosomiasis endemic states (Ondo and Kebbi).

Using three adapted versions of the screening tool against colposcopy and unit costs of activities, performance of the tool and cost-effectiveness analysis were evaluated. Compared to the colposcopy, the performance of the FGS screening tool varied using different screening definitions. When only direct and indirect contact with surface water was considered it demonstrated a sensitivity of 66.7% (95%CI: 60.1-72.8) and specificity of 35.4% (30.2-40.9); and when water contact together with any self-reported urogenital symptom was considered, the sensitivity and specificity were 50.2% (95%CI: 43.5 - 56.9) and 48.0% (95%CI: 42.5 - 53.6) respectively. Activity based micro-costing, showed that the FGS screening tool could screen individuals for approximately US$11 per woman, compared with US$20 per woman when using colposcopy. The screening tool detected far fewer true positive cases than the reference standard, assuming colposcopy did not miss true positives. The sub-optimal performance of the screening tool indicates the need for further refinement, to balance cost and effectiveness.

## Introduction

Living with a gynaecological condition that is poorly understood by many healthcare providers, rarely diagnosed and inadequately managed within the local healthcare system, can have detrimental consequences for individual health. In many schistosomiasis-endemic communities in sub-Saharan Africa, this is the reality for millions of women and girls affected by female genital schistosomiasis (FGS), a neglected gynaecological disease caused by *Schistosoma* [1, 2].

FGS disproportionately affects women and girls in the poorest communities, who rely on unsafe water sources such as rivers or ponds for domestic use and therefore bear the greatest risk of *Schistosoma* infection [1, 3]. Several studies across sub-Saharan Africa, including Nigeria, estimate that the burden of FGS ranges between 23% to 70% [1, 3, 4].

FGS can be treated with a single dose of praziquantel (40mg/kg), which kills the *Schistosoma* worms and stops progression of the disease. Although treatment with praziquantel is effective at eliminating parasites, it does not repair existing scars [5, 6], thus underscoring the need for early detection and treatment to prevent irreversible damage [7].

FGS remains largely invisible in clinical practice and is often misdiagnosed as sexually transmitted infections (STIs) due to similarity of symptoms [8–10]. This invisibility is due to a broad range of systemic factors present in many low-income countries (LICs): limited healthcare infrastructure, poor knowledge of the disease among communities, the absence of FGS topics in medical curricula, and the prohibitive cost of colposcopy, the procedure which the World Health Organization (WHO) recommends for visual confirmation of FGS [11, 12]

Colposcopy examination of the cervix is costly, resource intensive and requires specialist skills which are not available in primary healthcare facilities in endemic settings [13–15]. Molecular diagnostics, such as polymerase chain reaction (PCR), recombinase polymerase amplification (RPA), and loop-mediated isothermal amplification (LAMP) can detect *Schistosoma* deoxyribonucleic acid (DNA) from cervicovaginal lavage, vaginal self-swab, or cervical self-swab samples to identify FGS. However, this approach remains largely impractical for routine use in most endemic countries [16–18] due to its dependence on laboratory infrastructure, cold-chain management, and technical expertise, which are neither available nor affordable in most endemic settings [19, 20]. Emerging evidence indicates that self-sampling approaches and point-of-care molecular assays may help reduce both per-test costs and the need for facility-based clinical examinations by enabling community-based sample collection and simplified processing workflows [17, 21]. However, the evidence base of this approach remains limited.

In the absence of feasible ways to detect FGS in rural schistosomiasis endemic communities, where majority of affected women live, simple screening tools based on the exposure to schistosomiasis risk and FGS self-reported symptoms have emerged as a potential alternative. One such tool was developed by the Calling Time on Neglected Tropical Diseases (COUNTDOWN) research programme funded by UK AID [22] that was designed as an easy-to-use, non-technical method for healthcare workers in primary care settings in 2021 [23].

While simple FGS screening tools promise affordability and scalability of the disease detection and management, there are concerns about their sensitivity and specificity [24]. Evaluating the specificity and sensitivity of screening tools is critical, as these measures are key indicators of diagnostic accuracy and determine whether a tool is appropriate for wider use [25].

Furthermore, while effectiveness remains a primary consideration when evaluating any screening method or tool, the associated costs and resource requirements are also critical for wider adoption and use. Cost considerations are particularly essential for FGS screening, as the current diagnostic techniques are expensive and logistically difficult and the evidence on the use and economic value of alternative approaches is important but remains very limited.

In this paper, we present findings from evaluating the performance of adapted versions of the COUNTDOWN screening tool against colposcopy examination. We assess screening results based on three groups of questions used as thresholds for FGS risk and estimate the costs and cost-effectiveness of each approach versus colposcopy. We also examine the financial implications of implementing each approach, recognizing that cost is a decisive factor for health policy and intervention design.

## Materials and Methods

### Study design and settings

The COUNTDOWN tool and colposcopy were applied in a self-selected sample of women aged 15 – 49 years living in schistosomiasis endemic communities in Ondo and Kebbi States, Nigeria. The study took place between April and July 2023. Detailed description of study design and settings have been previously described by Osinoiki et al (2025) [1, 3].

### Study sampling

Study participants were selected following a multi-staged approach. First, the two states were selected in consultation with the Federal Ministry of Health and Social Welfare (FMoHSW) based on the endemicity of schistosomiasis, accessibility and security considerations. Second, within each state, two Local Government Areas (LGAs) with a schistosomiasis prevalence >20% among school aged children were selected - Oluji/OkeIgbo and Irele in Ondo, and Argungu and Ngaski in Kebbi. Thirdly, within each LGA, five communities (villages) with moderate to high schistosomiasis endemicity were selected. Women aged 15-49 living in the selected villages were invited to come to the local primary healthcare facility using community mobilisation campaign approaches and town announcers. All women who volunteered to take part in the study and met the age criteria were included in the sample. In total, 919 women completed the screening questionnaire. The same number of women underwent colposcopy examinations. However, 264 participants from Irele LGA were excluded from the analysis as there were concerns about the quality of examination due to the unavailability of a trained gynaecologist in this LGA. Additionally, 94 women did not have available colposcopy results, and 11 women had missing responses to one or more of the questionnaire items used in the analysis. Therefore, the sensitivity and specificity of screening versus colposcopy were assessed based on the records of 550 participants. Details on the sampling procedure are presented elsewhere [3].

### Data Collection

#### FGS screening tool

The adapted COUNTDOWN tool is a short questionnaire, which contains ten questions asking women about their exposure to open water sources as well as the presence and severity of different FGS symptoms. These include vaginal discharge, bleeding after intercourse or spotting, genital itching or burning sensation, pelvic pain (lower abdominal pain) or pain during or after intercourse. For this study, we assessed the accuracy of FGS screening based on three combinations of questions included in the tool and compared each option with colposcopy findings, which served as the reference standard. Results from screening questionnaire and colposcopy were independently collected by research assistants and gynaecologists respectively. to the performers/readers of the index test. In option 1 (Definition 1), participants were classified as suspected FGS cases solely based on self-reported contact with surface water, such as rivers or streams. In option 2 (Definition 2), women were considered suspected cases if they reported surface water contact together with at least one severe self-reported urogenital symptom. In option 3 (Definition 3), women were classified as suspected cases if they reported surface water contact and any self-reported urogenital symptom, irrespective of symptom severity. All women were interviewed face to face by trained female research assistants using the Commcare based mobile application.

#### Clinical examination

Gynaecological examinations were performed by female gynaecologists using an ELM-2000 portable colposcope with 18× optical zoom lens and display magnification of 1–54× (ECOMED, Shenzhen, China). Women were defined as FGS positive if they have any of the FGS associated symptoms stated in the WHO FGS Pocket Atlas [19]. These include homogeneous sandy patches, grainy patches, rubbery papules or abnormal blood vessels with other lesions were present. Detailed procedure has been described by Osinoiki et al [3]. The cervico-vaginal surfaces were inspected using guidelines outlined in the WHO FGS Pocket Atlas [26].

#### Costing methodology and economic evaluation

Costs were collected and computed using a mixed method of top-down and bottom-up micro-costing [27]. Transaction lists were obtained from the key programme implementer’s accounting system (Sightsavers) and used for the top-down approach. However, cost disaggregation was not possible in some LGAs (mostly in Ondo state) which meant the top-down approach could not be used. Accordingly, additional information was obtained through consultations with the project manager and coordinators involved in the study, and costs were calculated using a bottom-up method (details Appendix Table 5).

Cost items were allocated to activities and then to screening interventions, when directly attributable (e.g. cost of a colposcope rental was directly attributable to colposcopy). When an item or activity was shared, its cost was evenly distributed across screening interventions (e.g. cost of vehicles were shared between colposcopy and questionnaire collection as gynaecologist, nurses and data collectors use the same vehicle to join the facility) (Figure 1). For more details on activities included see Appendix Table 6.

**Figure 1:**
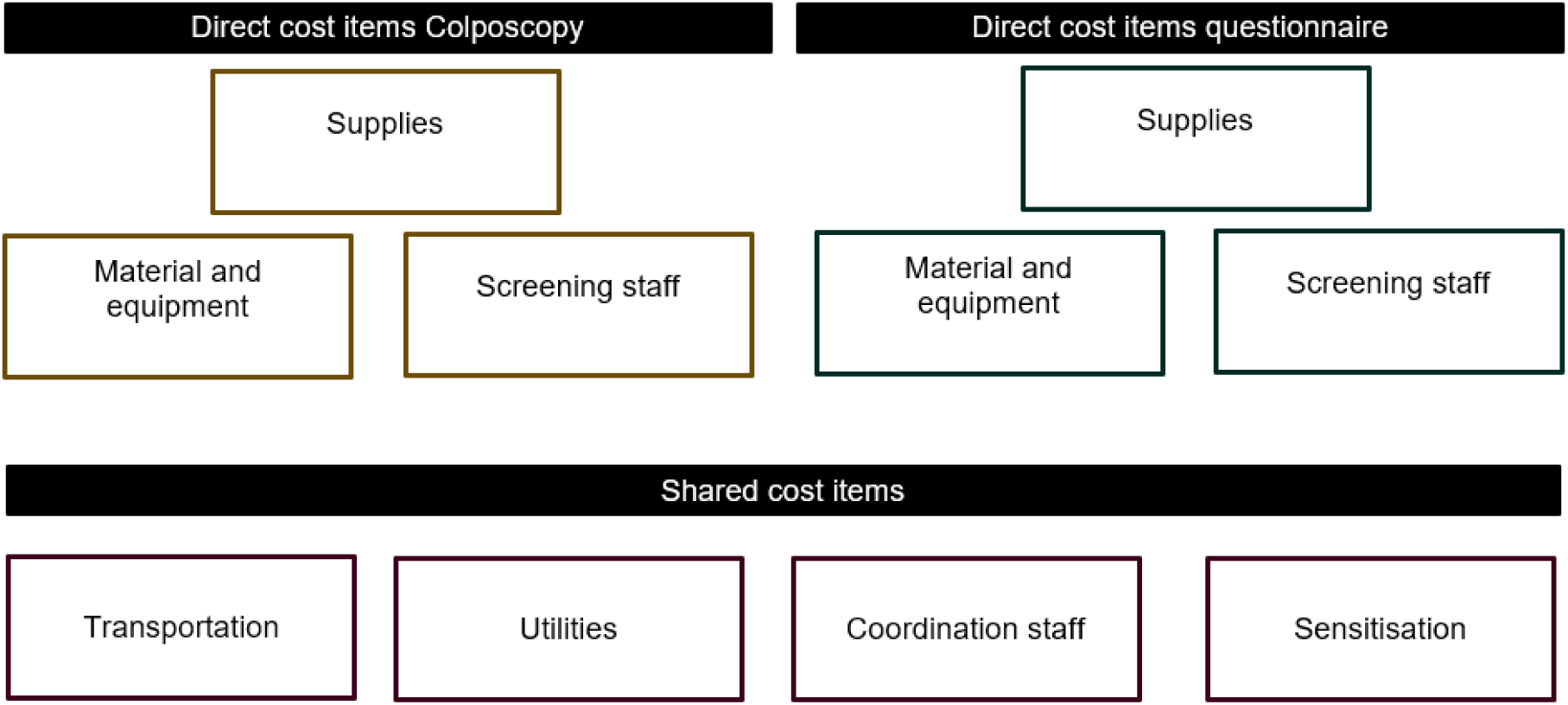
Items and activities allocation.

The analysis was conducted from the health provider perspective, including the Ministry of Health and Sightsavers, and the time horizon was restricted to the immediate screening intervention period.

Material and equipment costs were annualised using Turner et al (2023) methods, with a discount rate of 3% and a lifespan of five years [28]. All costs incurred were recorded in Nigerian Naira and converted to US$ 2023 using average monthly exchange rates [29].

Using the screening tool, the clinical examination data and unit costs calculated as described above, a cost-effectiveness analysis was performed. Four screening strategies were evaluated: Colposcopy-based screening (reference standard), FGS screening tool using Definition 1, FGS screening tool using Definition 2, and FGS screening tool using Definition 3 (more details in sub-section “clinical examination”). Colposcopy was treated as the diagnostic reference standard and served as the comparator for the cost-effectiveness calculations. Since treatment costs were not included (due to a lack of data) and since the effectiveness of praziquantel in reversing established FGS lesions remain unproven, effectiveness was not expressed in Disability-Adjusted Life Years averted or Quality-Adjusted Life Years gained [30]. Instead, the primary effectiveness outcome was the number of true positive FGS cases detected by each strategy (assuming perfect screening results for colposcopy). The first analysis consisted of comparing the cost per primary outcome by dividing the total cost of the screening intervention by the number of true positive cases, using colposcopy as reference standard (i.e. all colposcopy positive cases identified were assumed true positive). We applied a single unit cost for the screening tool across all three case definitions, as these were derived from the same questionnaire. The cost per true positive case was calculated by dividing the annual cost per woman screened by the number of true positive cases detected under each definition. As the screening options were considered as lower cost but potentially lower effectiveness alternatives to colposcopy, we additionally calculated decremental cost-effectiveness ratios (d-CER). d-CER is a metric that represents the cost saved for each unit of health outcome lost when an intervention is less effective, but also less costly than the comparator [31]. These were calculated using the following formula:

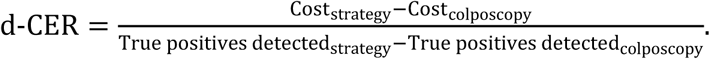

As these alternatives were less effective and less costly, the resulting d-CERs represent decremental cost-effectiveness ratios, reflecting the cost savings associated with detecting fewer true positive cases relative to colposcopy.

This simplified CEA did not incorporate long-term consequences of FGS, including increased HIV susceptibility, cervical precancer or cancer, or reproductive health complications. As mentioned above, the analysis also assumes that colposcopy as the gold reference standard despite known limitations in sensitivity and specificity [30].

#### Data analysis

Data was analysed using STATA 16.1 (Stata Corporation, College Station, TX) statistical package, R version 4.4.3, and Microsoft Excel (Microsoft Corporation, Redmond, WA, USA) [32, 33].

#### Specificity and sensitivity analysis

Descriptive statistics were presented using frequencies and cross tabulations. Specificity and sensitivity are reported overall and per state for each of the screening definitions using colposcopy results as a reference standard.

#### Ethical consideration

This study was approved by the National Health Research Ethics Committee of the Federal Ministry of Health and Social Welfare, Nigeria (reference: NHREC/01/01/2007) and by the State Research Ethics Committees in Kebbi Health Research Ethics Committee (KSHREC 107:033/2023) and Ondo Health Research Ethics Committee (OSHREC 08/05/2023/556). Informed written and oral consent was obtained from all participants for parasitological and gynaecological examinations. For participants <18 y of age, parents or guardians gave informed permission, and assent was obtained from the participants. Privacy and confidentiality of medical information were protected during and after the study.

Written (or thumbprint) informed consent was obtained from each eligible participant prior to data collection. For participants who could not read and write, information about the study was read out by study enumerators. All women were treated with a single dose of praziquantel (40 mg/kg) free of charge. Those who were diagnosed with other gynaecological conditions were referred to upper-level facilities for further investigation and treatment.

## Results

### Positive FGS results using the COUNTDOWN screening tool and colposcopy

Results for both screening and colposcopy were available for 550 women (358 in Kebbi, 192 in Ondo). Using the FGS screening tool, 65.5% (n=360), 22.7% (n=125) and 51.3% (n=282) were categorized as suspected of FGS according to Definitions 1, 2 and 3 respectively (Table 1). In Ondo state communities, the results were 77.6% (n=149), 13.0% (n=25) and 50.5% (n=97), respectively; and in Kebbi state communities, the results were 58.9% (n=211), 27.9% (n=100) and 51.7% (n=185), respectively.

**Table 1:**
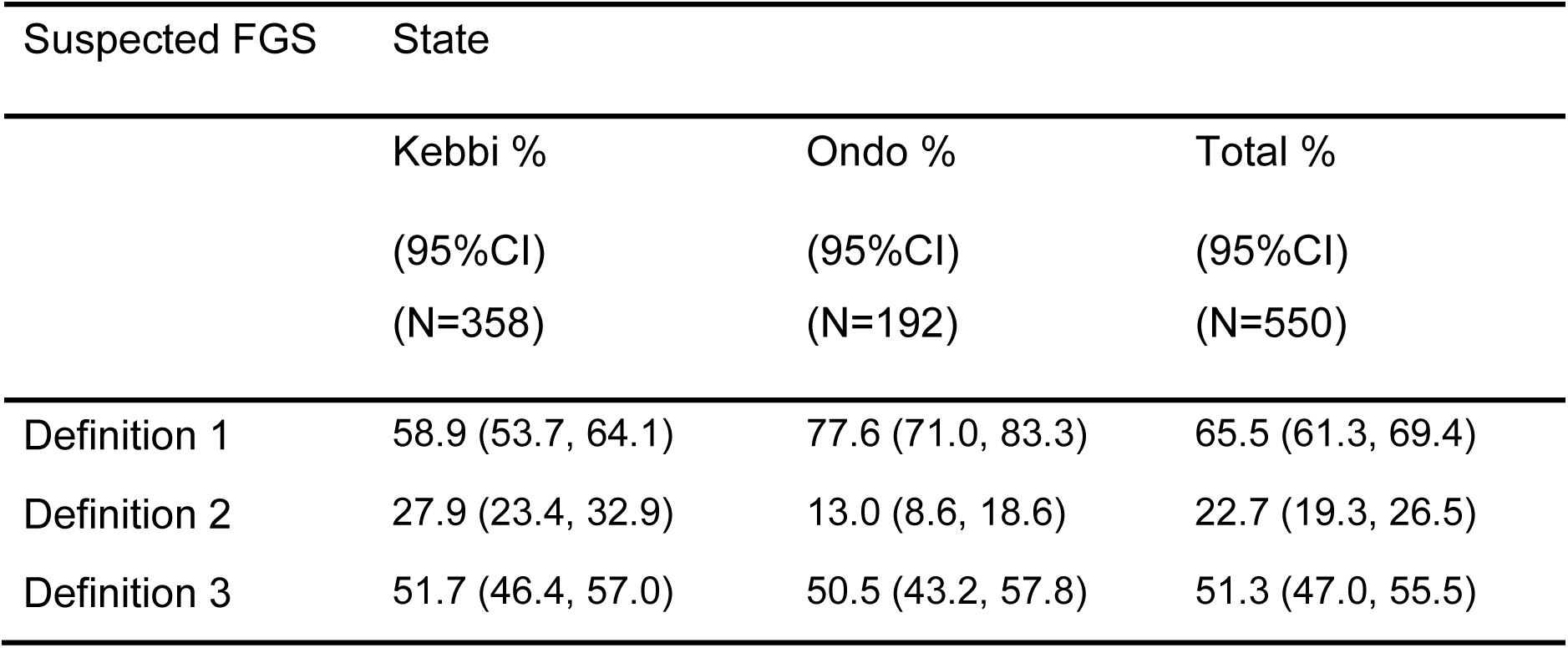
Proportion of women identified as suspected of FGS using the FGS screening tool definitions. Based on the colposcopy examination 40.9% of women overall (n=225), 34.4% (n=123) in Kebbi and 53.1% (n=102) in Ondo were diagnosed with FGS.

### Comparison of the FGS screening tool and colposcopy

Sensitivity, specificity, positive predictive value (PPV) and negative predictive value (NPV) for the three screening definitions are shown in Figure 1. The overall sensitivity and specificity of the screening tool using Definition 1 was 66.7% (95%CI 60.1, 72.8) and 35.4% (30.2, 40.9), respectively. The overall sensitivity and specificity of the screening tool using Definition 2 was 19.6% (14.6, 25.4) and 75.1% (70.0, 79.7) respectively. The overall sensitivity and specificity using Definition 3 was 50.2% (43.5, 56.9) and 48.0% (42.5, 53.6) respectively. Sensitivity and specificity varied markedly across sites. Overall, the tool performed better in Ondo communities and the tools sensitivity was the highest using Definition 1 (86.3%), but the tools specificity was the highest using Definition 2 (93.3%).

The overall positive predictive value (PPV) was relatively low across all definitions, varying from 35.2% for Definition 2 to 41.7% for Definition 1. PPV varied substantially across sites, being consistently higher for Ondo than Kebbi. Negative predictive value (NPV) varied overall from 57.4 % for Definition 2 to 60.5% for Definition 1 and showed less variation across sites (Figure1 and S1,S2, S3 and S4).

### Costing of screening tool and colposcopy

The micro-costing analysis, as shown in figures 2, 3 and 4, demonstrates clear differences in the cost structure of colposcopy versus questionnaire-based screening across Kebbi and Ondo states. Screening interventions were implemented sequentially in the same facilities, using the same transportation and sensitisation activities, utilities and coordination staff costs for both screening interventions. However, a difference was observed at the state level, where the total of shared costs (transportation, sensitisation, utilities and coordination staff) reached US$7 per woman screened in Kebbi and US$9.7 per woman screened in Ondo.

**Figure 2:**
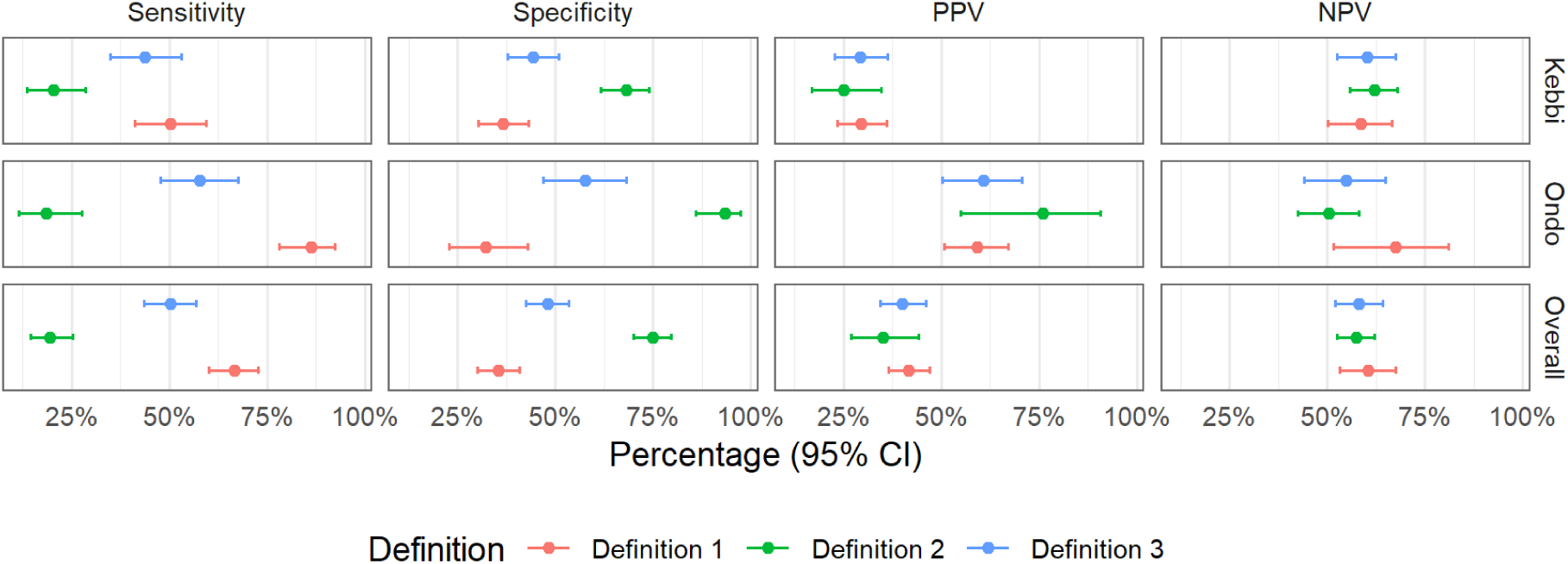
Sensitivity, specificity, PPV and NPV of the three FGS screening tool definitions, using colposcopy as reference standard – overall and by state. \***Abbreviations**: PPV-Positive Predictive Value, NPV-Negative Predictive Value, CI-Confidence Interval

**Figure 3:**
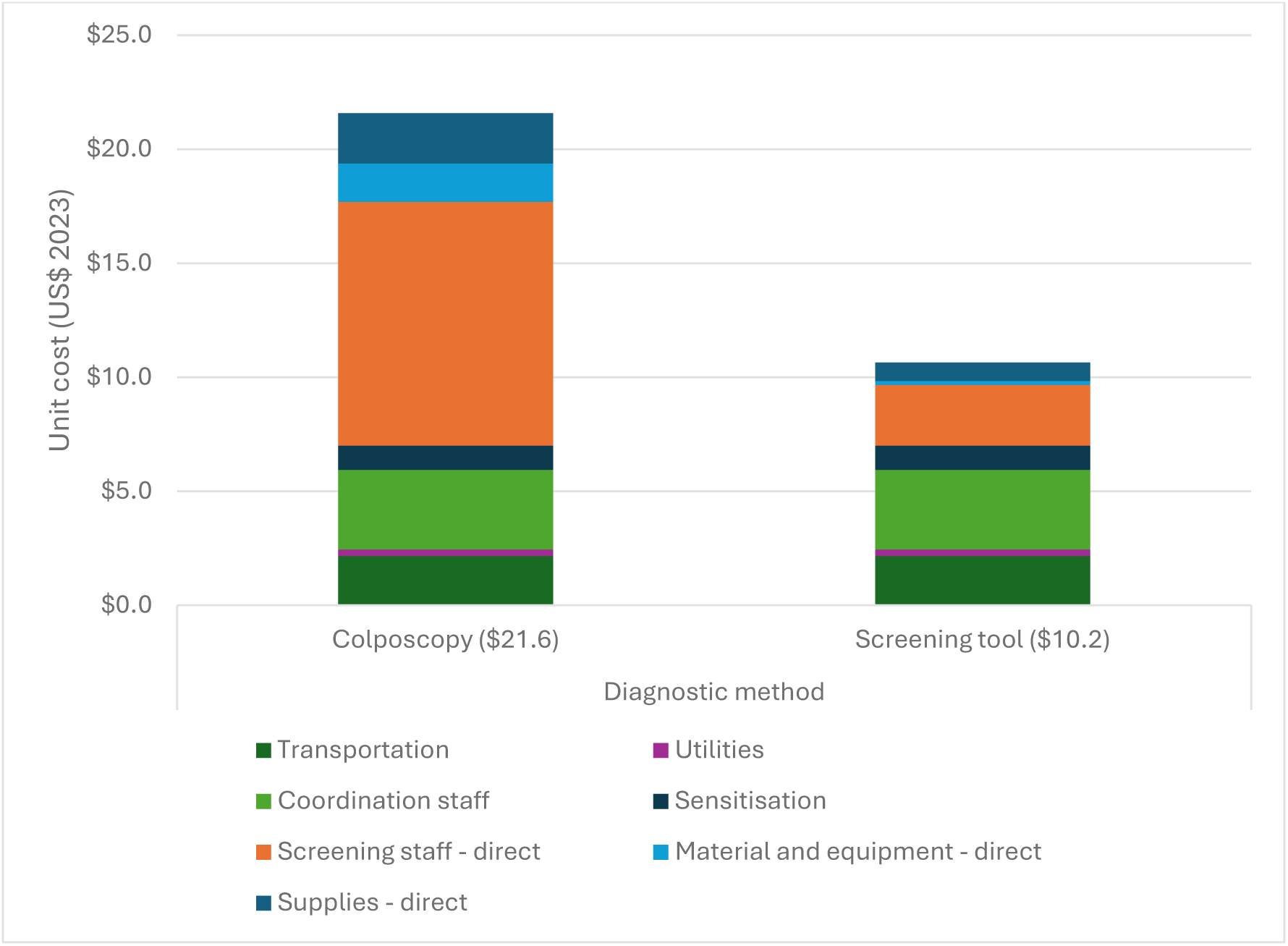
Unit cost per screening methodology in Kebbi, n=358 (in US$ 2023)

**Figure 4:**
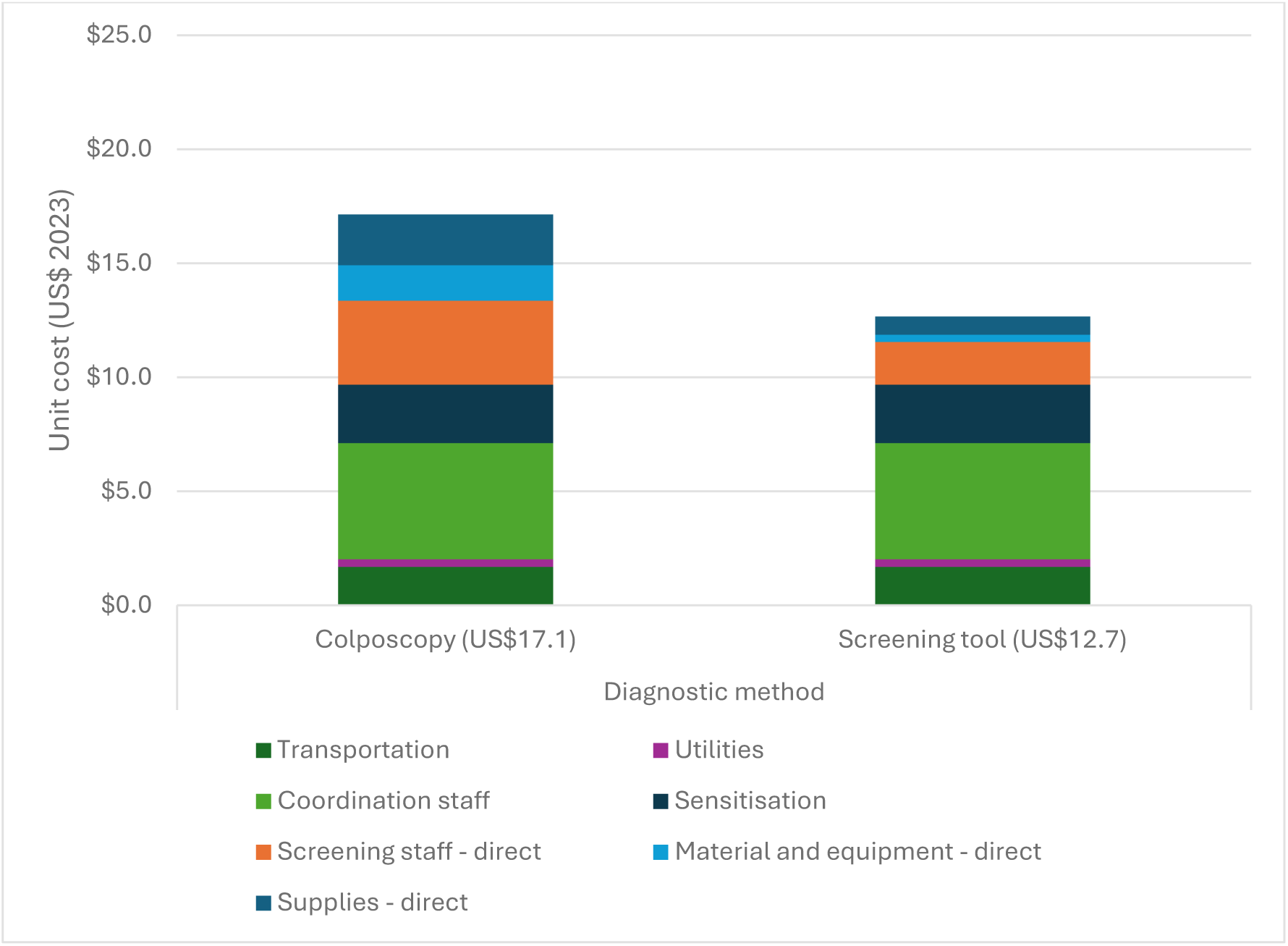
Unit cost per screening methodology in Ondo, n=192 (in US$ 2023)

**Figure 5:**
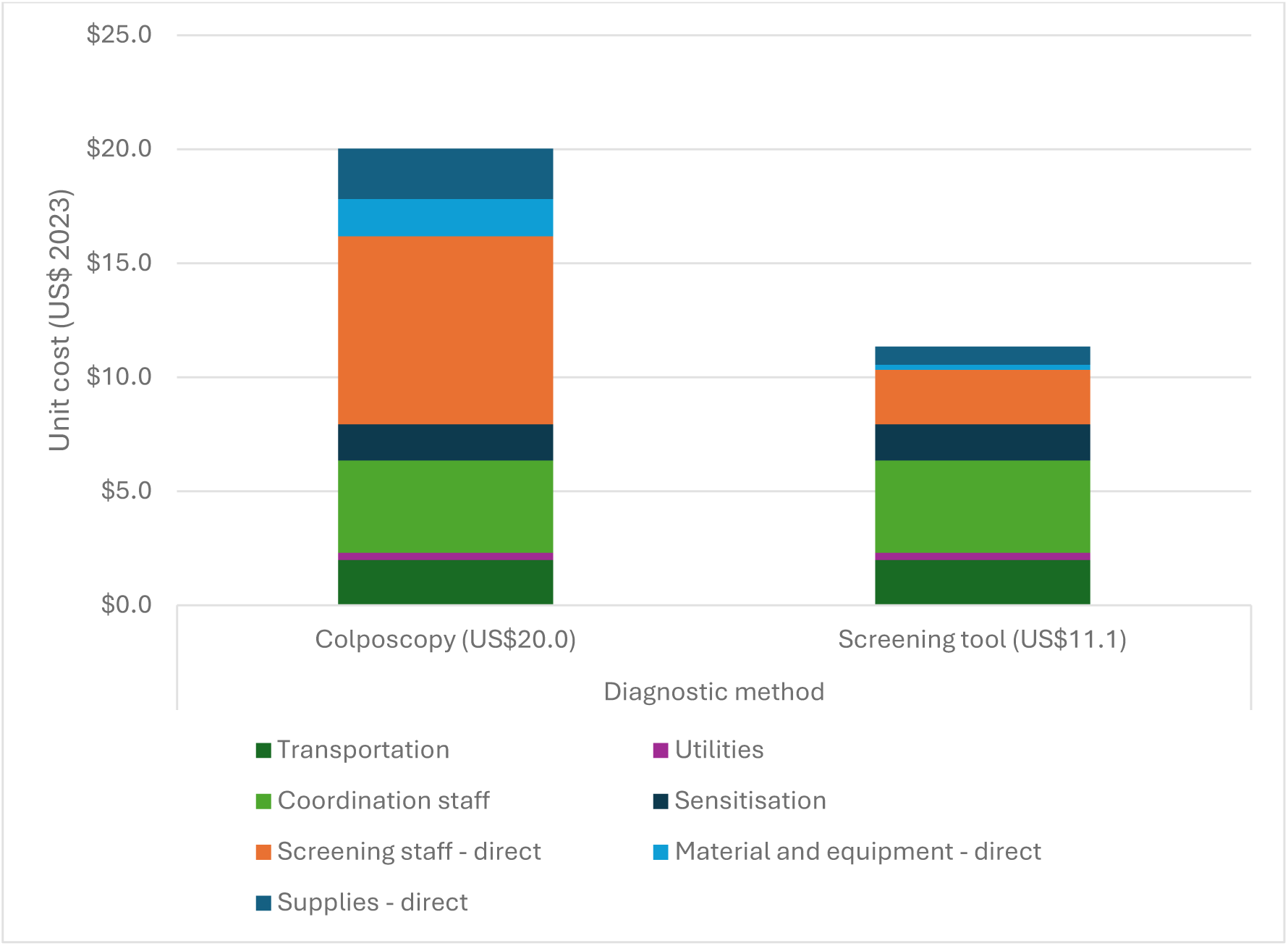
Unit cost per screening methodology overall, n=550 (in US$ 2023)

In terms of total cost of the screening intervention, colposcopy cost US$21.6 per woman screened in Kebbi, compared with US$10.2 for questionnaire screening. In Ondo the corresponding costs were lower for colposcopy (US$17.1 per woman screened), but higher for screening tools (US$12.7) (figure 2 and 3).

Across all settings, personnel costs for screening constituted the largest cost component for colposcopy, accounting for US$10.7 in Kebbi and US$3.7 in Ondo (figures 2 and 3). In contrast, questionnaire-based screening incurred substantially lower personnel screening costs (US$2.7 per woman screened in Kebbi and US$1.9 in Ondo). Only one data collector was needed to interview each patient, whereas a gynaecologist and a nurse were hired for the colposcopy. Supplies contributed modestly but consistently to total costs in both approaches across states, with supplies fixed at US$2.2 for colposcopy and US$0.8 for questionnaires. The colposcopy procedure required more medical supplies, such as disposable speculum, absorbent under pads or acetic acid. Colposcopy also required more material and equipment than screening tool interviews (US$0.2 per woman screened in Kebbi and US$0.3 in Ondo), where colposcope were rented as well as gynaecological couch, for a unit cost of US$1.7 per women screened in Kebbi and US$1.6 in Ondo. Overall, the findings indicate that the higher cost of colposcopy is primarily driven by personnel-intensive service delivery and greater supplies and equipment needed.

In Kebbi, the cost per true positive case reached US$63 for colposcopy and US$61 for Definition 1 of the screening tool, increasing to US$152 for Definition 2 and US$201 for Definition 3 (table 2). In Ondo, costs were lower overall, with US$32 for colposcopy and US$28 for Definition 1, then rising to US$128 for Definition 2 and US$41 for Definition 3. Overall, the cost per true positive case was US$49 for colposcopy, US$42 for Definition 1, compared with US$142 for Definition 2 and US$80 for Definition 3.

**Table 2:** Cost per FGS screening methodology per true positive case by state (US$ 2023)

| **Kebbi (n=358)** | **Colposcopy** | **Def. 1** | **Def. 2** | **Def. 3** |
| --- | --- | --- | --- | --- |
| Cost per woman screened | US\$22 | US\$11 | US\$11 | US\$11 |
| Positive case identified | 123 | 211 | 100 | 185 |
| True positive case | Reference | 62 | 25 | 19 |
| Cost per true positive case | US\$63 | US\$61 | US\$152 | US\$201 |
| d-CER | Reference | US\$40 | US\$25 | US\$24 |
| **Ondo (n=192)** | **Colposcopy** | **Def. 1** | **Def. 2** | **Def. 3** |
| Cost per woman screened | US\$17 | US\$13 | US\$13 | US\$13 |
| Positive case | 102 | 149 | 25 | 95 |
| True positive case | Reference | 88 | 19 | 59 |
| Cost per true positive case | US\$32 | US\$28 | US\$128 | US\$41 |
| d-CER | Reference | US\$61 | US\$10 | US\$20 |
| **Overall (n=550)** | **Colposcopy** | **Def. 1** | **Def. 2** | **Def. 3** |
| Cost per woman screened | US\$20 | US\$11 | US\$11 | US\$11 |
| Positive case | 225 | 360 | 125 | 280 |
| True positive case | Reference | 150 | 44 | 78 |
| Cost per true positive case | US\$49 | US\$42 | US\$142 | US\$80 |
| d-CER | Reference | US\$64 | US\$26 | US\$32 |

It should be noted that the cost per true positive does not account for missed true positive cases. Across both sites, the screening tool reduced costs but at the expense of lower diagnostic sensitivity, resulting in missed cases. This trade-off is reflected in the d-CER, expressed as cost savings per true positive forgone, which ranged from US$24 to US$40 in Kebbi and US$20 to US$61 in Ondo.

## Discussion

This study examined the performance and financial implications of a simple questionnaire-based FGS screening tool to detect FGS cases as an alternative to more expensive and logistically complicated colposcopy in schistosomiasis-endemic communities in Nigeria. Across both states, the proportion of individuals classified as suspected FGS cases varied depending on the definition used. Definition 1 which relied solely on reported surface-water contacts identified the highest proportion of suspected cases (over 65% overall), compared to the definitions based on a combination of surface water contact and urogenital symptoms (Definitions 2 and 3). Findings from the test performance analysis showed that all three screening definitions applied independently demonstrated suboptimal sensitivity and specificity, although there were regional variations with the tool performing considerably better in Ondo than in Kebbi communities. The prevalence of FGS on colposcopy was higher in Ondo areas. Therefore, a better performance of the tool, particularly in higher PPV, is expected.

These results indicate that the tool (as applied in this study) risks misclassifying a considerable number of women as FGS positive, which can be problematic if the screening is used for deciding on the eligibility for praziquantel treatment. For example, asking women about their access to open water sources showed the highest sensitivity (around 67% overall and over 86% in Ondo), but even with this broad question, around one third of women (14% in Ondo) diagnosed with FGS by colposcopy will be missed. Moreover, the specificity of this screening approach was low (35%), which means that two thirds of women who do not have FGS by colposcopy will be classified as eligible for praziquantel. In addition, high numbers of false positives may strain referral systems and generate unnecessary anxiety for affected women.

Combining self-reported urogenital symptoms with the source of water criterion improved only specificity of screening, particularly when the severity of symptoms was accounted for (75% overall and over 93% in Ondo). Our findings are different from the study conducted in Tanzania where the symptom-based screening checklist demonstrated high sensitivity of over 85% but very low specificity of less than 9% [34]. The differences may be due to the exclusion of three questions in the screening tool used in our study, and the use of a digital colposcope as a reference in the Tanzania study. Another important distinction is that the screening tools previously tested elsewhere typically applied a graded scoring system, allowing risk to be categorised by severity. In contrast, our adaptation applied simplified binary classifications without symptom weighting, which may have reduced the accuracy of categorization [29]. This reinforces the need for careful adaptation and validation of screening tools, ensuring that modifications do not unintentionally compromise the performance.

In addition, while water contact is a known risk factor for schistosomiasis [1, 3, 35], reliance on a single self-reported exposure variable may be vulnerable to recall bias, especially in the communities where water infrastructure and access to portable water has improved over time [9, 36]. Self-reported urinary and gynaecological symptoms are also prone to recall bias [37], which may also affect the sensitivity and specificity of screening. The inclusion of non-specific FGS-related symptoms such as genital itching, discharge, or painful intercourse into the tool, may also affect the accuracy of screening, as these symptoms may be relevant to sexual or reproductive health conditions rather than FGS [38–40]. Consequently, while the idea of identifying a simple, low-cost screening tool that can be administered by primary healthcare workers without specialist equipment and close to local communities is very appealing, our findings show that this is not a straightforward task. Our economic evaluation does not produce strong evidence to support this approach to triage patients for treatment either. Although the unit costs of screening were lower than those of colposcopy, the cost per correctly identified case was higher for all screening definitions, reflecting the reduced number of true positive cases detected. This trade-off is further illustrated by the positive decremental cost-effectiveness ratios, indicating that cost savings are achieved at the expense of missed true cases (Table 2), thereby limiting the effectiveness and economic value of these alternative definitions.

Our activity-based costing analysis showed that the unit cost of implementing the FGS screening tool was US$11 per woman screened, while the cost of colposcopy was US$20. It is difficult to compare these results as there are very few economic evaluations of FGS interventions, but our colposcopy costs appear to be comparable with other studies, after adjusting to inflation and converting to US$ 2023, which estimated the direct cost of conducting a colposcopy in a clinic at US$9.6 per examination in Uganda, US$13.4 in India and US$20.7 in Nicaragua [41]. The only other study which has assessed the cost-effectiveness of alternative screening approaches [30] evaluated the costs of home-based self-sampling and clinic-based sampling of genital swabs in Zambia at US$7.7 and US$ 11 respectively (in US$ 2021), which cannot be compared with the screening tool used in our study.

As stated earlier, the relatively high cost of the questionnaire-based screening (US$11 per woman screened) in our study was largely driven by the costs of coordination, hiring and transporting external data collectors. If integrated in the local service delivery systems and conducted by facility personnel, the costs would be considerably reduced, although the logistics of the tool administration (e.g. printing and storage of data collection forms) needs to be considered. Bringing these additional logistical complexities, without considerable added value of the questionnaire for treatment decision-making, cannot be justified. In fact, considering the screening results and context, presumptive treatment of women of reproductive age using praziquantel, would be the most effective strategy in high endemic communities, as supported by other studies [42]. In our research, treating directly all 550 women, using an internally estimated unit cost of US$0.47 per dose, would amount to US$259, which is significantly less expensive than either the screening or colposcopy. It is however important to note that evidence on the effectiveness of praziquantel once FGS has developed remains limited. Therefore, despite presumptive treatment, detection and appropriate management of FGS should continue to address FGS-related lesions and other associated conditions [43, 44]. Moreover, supply of praziquantel in large quantities has become difficult in the past few years, as some manufacturers discontinued the production due to high manufacturing costs and low purchasing power in the affected low-income country markets.

In this situation, improving the performance of the screening tool, which could be used for both prioritising patients for treatment and referral to upper-level facilities for further diagnosis and management will be of critical importance. The tool may also be useful for training primary healthcare workers and increasing their knowledge about risks and symptoms of FGS. They can then use this knowledge and tool in health education campaigns, signposting women to services for further investigation and treatment, particularly in settings where colposcopy services cannot be easily reached.

Moreover, our analysis did not account for comorbidities associated with FGS or the consequences of missed diagnoses, both of which represent important public health issues. As highlighted by Lamberti et al. and Webb et al., FGS is linked to an increased risk of HIV contraction and cervical precancer, meaning that leaving the disease undetected may have substantial implications for women’s health [30, 42]. Undetected cases may progress to severe disease, including cervical malignancy, and ultimately contribute to preventable morbidity and mortality. These trade-offs highlight the need for a balanced policy approach and further research to consider costs associated with integrated diagnostics (e.g. FGS and cervical cancer).

The economic cost of interventions is a critical determinant in the adoption and integration of screening and diagnostic methods. Building on the cost analysis, the simplified cost-effectiveness assessment results showed that while the FGS questionnaire-based screening was implemented at lower costs than colposcopy, it also missed a large proportion of true positive cases. Indeed, in the best case the maximum d-CER is US$64 saved for each missed case, beyond ethical importance, it is unlikely that these savings outweigh the health benefits associated with correctly identifying and treating infected women. Colposcopy therefore remained the most effective strategy, identifying 225 positives across the settings, assuming perfect sensitivity and specificity (Table 1). At present, primary healthcare facilities in Nigeria do not have access to the equipment and skills required for colposcopy and examinations can be performed only through periodic outreach services, which cannot be available at scale at US$20 per examination. Future research is needed to look at the potential to reduce colposcopy costs and the effectiveness and cost-effectiveness of using the screening tool to triage patients for more complex diagnostics using colposcopy or molecular assays. For example, Definition 1 option tested in this study may offer a pragmatic first screening step enabling primary health workers identify women for more complex confirmatory diagnosis. However, this approach would still leave a substantial proportion of women undetected if used in isolation, underscoring the need for further refinement of the tool. Also, integrating FGS screening with cervical cancer screening or in sexual reproductive health services could potentially further increase opportunities for case identification and treatment and possibly optimising resources [19].

This study has limitations that should be considered when interpreting the results. Firstly, the sample size was relatively small and based on self-selection, which may not be representative of the broader population. Secondly, a few questions from the original screening tool were excluded in this study and there was no validation of the screening tool in local languages. To improve the generalizability of our findings, we also need to expand our evaluation by scoring the risks and symptoms. Thirdly, this study does not account for potential long-term savings from early detection and treatment of FGS, which could underestimate the overall cost-effectiveness of the screening interventions. Finally, the cost-effectiveness comparison assumes that colposcopy is a gold standard whereas this screening method is not perfectly sensitive and specific.

## Conclusion

To our knowledge, this is the first assessment on the performance of FGS screening tool and cost-effectiveness in Nigeria. It provides one of the first combined assessments of the screening tool performance and costs. Although, the screening definitions tested here did not demonstrate high sensitivity or specificity to serve as stand-alone case detection methods, our findings highlight the practical value of simple algorithms in settings where colposcopy services are scarce or difficult to sustain. Despite their limitations, water contact and self-reported urogenital symptoms remain important initial indicators that primary healthcare workers can use to identify women who may require further clinical evaluation.

At the same time, the results underscore the need to strengthen and refine screening approaches before they can be used at scale. Improving the structure of the tool, validating local adaptations, and exploring symptom weighting models could enhance accuracy. Given the significant resource constraints in endemic areas, an approach that uses low-cost screening for early identification, coupled with targeted referral for colposcopy or more advanced diagnostics, may offer the most feasible pathway for improving FGS detection and care. Continued investment in training, community awareness, and health system capacity will be essential to ensure that women and girls affected by FGS are not missed and receive timely, appropriate support.

## Funding: None

This work was supported by Sightsavers, United Kingdom. Sightsavers is the direct employer of authors (OO, GT, ASO, MI, AJ, IJ, RS and ES) and supported the time allocation for analysis of data used in this study.

## Data Availability

All data generated or analysed during this research (and its online supplemental files) will be available when all articles for publications from the research have been submitted. The datasets generated and/or analysed during the current study are not publicly available because other manuscripts are being prepared from the findings of this dataset. However, datasets are available from the corresponding author upon reasonable request.

## Appendix

**Table 3:** Sensitivity of the FGS screening tool, using different case definitions, and colposcopy as the reference standard, by study site and overall. Data are percentages [95% CI].

|  | Kebbi | Ondo | Overall |
| --- | --- | --- | --- |
| Definition 1 | 50.4 [41.3, 59.5] | 86.3 [78.0, 92.3] | 66.7 [60.1, 72.8] |
| Definition 2 | 20.3 [13.6, 28.5] | 18.6 [11.6, 27.6] | 19.6 [14.6, 25.34] |
| Definition 3 | 43.9 [35.0, 53.1] | 57.8 [47.67, 67.6] | 50.2 [43.5, 56.9] |

**Table 4:** Specificity of the FGS screening tool, using different case definitions, and colposcopy as the reference standard, by study site and overall. Data are percentages [95% CI].

|  | Kebbi | Ondo | Overall |
| --- | --- | --- | --- |
| Definition 1 | 36.6 [30.4, 43.1] | 32.2 [22.8, 42.9] | 35.4 [30.2, 40.9] |
| Definition 2 | 68.1 [61.7, 74.0] | 93.3 [86.1, 97.5] | 75.1 [70.0, 79.7] |
| Definition 3 | 44.3 [37.8, 50.9] | 57.8 [46.9, 68.1] | 48 .0 [42.5, 53.6] |

**Table 5:**
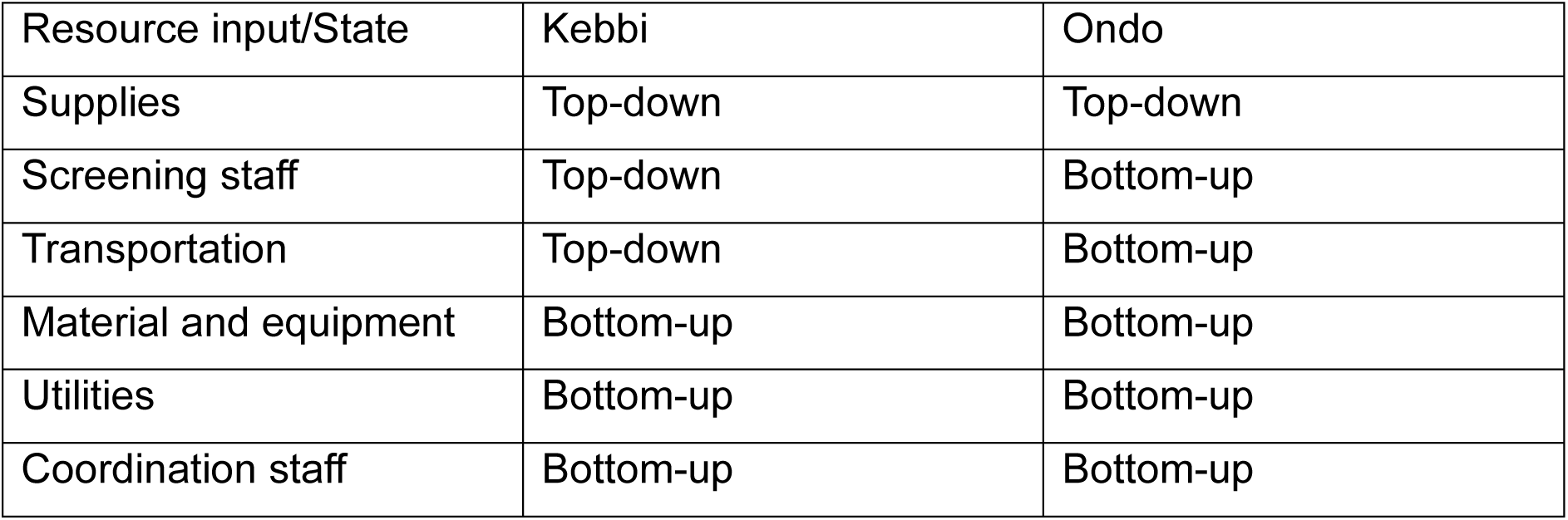
PPV of the FGS screening tool, using different case definitions, and colposcopy as the reference standard, by study site and overall. Data are percentages [95% CI].

**Table 5:**
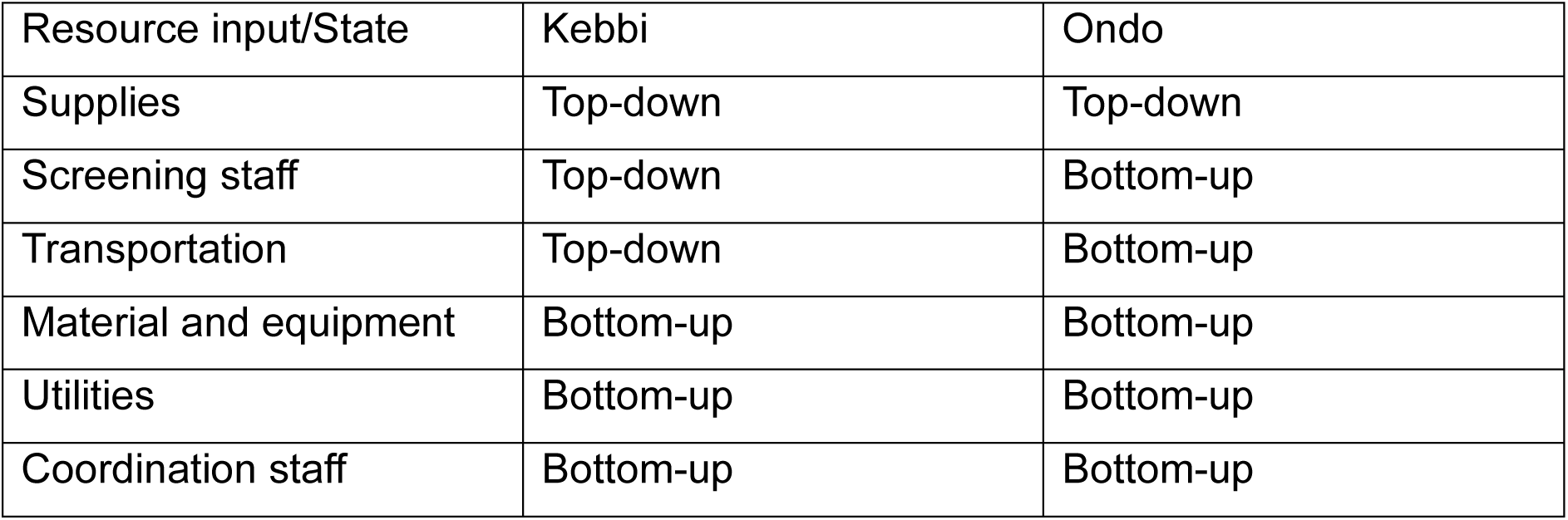
Costing method used by resource input and state.

**Table 6:** NPV of the FGS screening tool, using different case definitions, and colposcopy as the reference standard, by study site and overall. Data are percentages [95% CI].

|  | Kebbi | Ondo | Overall |
| --- | --- | --- | --- |
| Definition 1 | 58.5 [50.1, 66.6] | 67.4 [51.5, 80.9] | 60.5 [53.2, 67.5] |
| Definition 2 | 62.0 [55.8, 68.0] | 50.3 [42.5, 58.1] | 57.4 [52.6, 62.2] |
| Definition 3 | 60.1 [52.4, 67.5] | 54.7 [44.2, 65.0] | 58.2 [52.1, 64.2] |

**Table 6:** Items included and type of allocation.

| Activities | Items included | Type of allocation |
| --- | --- | --- |
| Supplies | Absorbent under pads, Acetic acid 2% in alcohol, Disposable Speculum, Saline, Wooden Spatula, Disposable laboratory coats, Disposable Masks, Hand Sanitiser, Latex Gloves, Liquid Soap, Tissue etc. | Direct |
| Screening staff | Gynaecologists, nurse assistant and data collectors | Direct |
| Material and equipment | Colposcope rental, Fuel to hire colposcope, Gynaecological couch | Direct |
| Transportation | Vehicle hire or purchased (annualised) and driver allowance | Shared |
| Utilities | Generator (fuel) at facility, printing stationary | Shared |
| Coordination staff | Daily allowance, per diem or salary for all staff required for the coordination and implementation of the pilot screening interventions: Research Coordinator, Research Associate, Research Adviser, Principal investigator, Scientific officer/team lead, Federal Ministry of Health Director, Zonal officers, State Coordinator, State Team 1 (Department of Public Health), State Team 2 (Primary Health Care Board ), State Team 3 (State Health Promotion Office), Local Government Areas for Neglected Tropical Diseases staff | Shared |
| Sensitisation | Town announcer allowance | Shared |

## Bibliography

1. Ekpo, U.F., et al., Female genital schistosomiasis (FGS) in Ogun State, Nigeria: a pilot survey on genital symptoms and clinical findings. Parasitology Open, 2017. 3: p. e10.

2 Umbelino-Walker, I., et al., Integration of female genital schistosomiasis into HIV/sexual and reproductive health and rights and neglected tropical diseases programmes and services: a scoping review. Sex Reprod Health Matters, 2023. 31(1): p. 2262882.

3. Osinoiki, O., et al., The burden and risk factors of female genital schistosomiasis in two schistosomiasis-endemic states of Nigeria. International Health, 2025. 17(Supplement_1): p. i73–i82.

4. Gyang, V.P., et al., Investigating outcomes of female genital schistosomiasis in communities in Ogun State, Nigeria: a pilot cross-sectional study. Transactions of The Royal Society of Tropical Medicine and Hygiene, 2025. 119(4): p. 367–374.

5. Anchang-Kimbi, J.K., et al., Diagnosis of female genital schistosomiasis and pre-emptive treatment with praziquantel: a community-based pilot intervention study in Tiko, Cameroon. Reprod Health, 2026. 23(1): p. 39.

6. Nemungadi, T.G., et al., Establishing and Integrating a Female Genital Schistosomiasis Control Programme into the Existing Health Care System. Trop Med Infect Dis, 2022. 7(11).

7. Oluwole, A.S., et al., Towards elimination of genital schistosomiasis in Africa: Outlining strategic public health objectives and measures to protect future generations. Parasitology, 2025. 152(14): p. 1554–1560.

8. Kukula, V.A., et al., A major hurdle in the elimination of urogenital schistosomiasis revealed: Identifying key gaps in knowledge and understanding of female genital schistosomiasis within communities and local health workers. PLOS Neglected Tropical Diseases, 2019. 13(3): p. e0007207.

9. Orish, V.N., et al., The parasitology of female genital schistosomiasis. Current Research in Parasitology & Vector-Borne Diseases, 2022. 2: p. 100093.

10. Rossi, B., et al., Female Genital Schistosomiasis: A Neglected among the Neglected Tropical Diseases. Microorganisms, 2024. 12(3): p. 458.

11. Sturt, A.S., et al., Beyond the barrier: Female Genital Schistosomiasis as a potential risk factor for HIV-1 acquisition. Acta Tropica, 2020. 209: p. 105524.

12. Jacobson, J., et al., Addressing a silent and neglected scourge in sexual and reproductive health in Sub-Saharan Africa by development of training competencies to improve prevention, diagnosis, and treatment of female genital schistosomiasis (FGS) for health workers. Reprod Health, 2022. 19(1): p. 20.

13. Bustinduy, A.L., et al., *Chapter One - An update on female and male genital schistosomiasis and a call to integrate efforts to escalate diagnosis, treatment and awareness in endemic and non-endemic settings: The time is now*, in Advances in Parasitology, D. Rollinson and R. Stothard, Editors. 2022, Academic Press. p. 1–44.

14. Fleming, F.M., et al., Now, more than ever, it’s time to address the neglect of female genital schistosomiasis. Parasitology, 2025. 152(14): p. 1570–1577.

15. Wasson, P.S., et al., Female Genital Schistosomiasis: Translational Challenges and Opportunities: Outputs and actions from a consultative, collaborative and translational workshop. Parasitology, 2025.

16. Galappaththi-Arachchige, H.N., et al., Evaluating diagnostic indicators of urogenital Schistosoma haematobium infection in young women: A cross sectional study in rural South Africa. PLOS ONE, 2018. 13(2): p. e0191459.

17. Archer, J., et al., Validation of the isothermal Schistosoma haematobium Recombinase Polymerase Amplification (RPA) assay, coupled with simplified sample preparation, for diagnosing female genital schistosomiasis using cervicovaginal lavage and vaginal self-swab samples. PLOS Neglected Tropical Diseases, 2022. 16(3): p. e0010276.

18. Lamberti, O., et al., Female genital schistosomiasis burden and risk factors in two endemic areas in Malawi nested in the Morbidity Operational Research for Bilharziasis Implementation Decisions (MORBID) cross-sectional study. PLOS Neglected Tropical Diseases, 2024. 18(5): p. e0012102.

19. Shanaube, K., et al., Zipime-WekaSchista study protocol: a longitudinal cohort study and economic evaluation of an integrated home-based approach for genital multipathogen screening in women, including female genital schistosomiasis, human papillomavirus, Trichomonas and HIV in Zambia. BMJ Open, 2024. 14: p. e080395.

20. Ajibola, O., et al., Tools for Detection of Schistosomiasis in Resource Limited Settings. Med Sci (Basel), 2018. 6(2).

21. Sturt, A.S., et al., Genital self-sampling compared with cervicovaginal lavage for the diagnosis of female genital schistosomiasis in Zambian women: The BILHIV study. PLoS Negl Trop Dis, 2020. 14(7): p. e0008337.

22. Consortium, C., Calling time on Neglected Tropical Diseases. 2026.

23. Sightsavers, L.S.o.T.M., Federal Ministry of Health of Nigeria, Federal University of Agriculture of Abeokuta, Federal Medical Centre of Abeokuta, Ministry of heealth of Ogun State, State Hospital Soeknu, Health Worker Training Guide for Managing Femal Genital Schistosomiasis (FGS) in Primary Health Care. 2021.

24. Rogers, E.Q., et al., Developing and validating a screening tool for female genital schistosomiasis in urban Zambia. Frontiers in Tropical Diseases, 2024. 4: p. 1308129.

25. Shreffler, J. and M.R. Huecker, Diagnostic testing accuracy: Sensitivity, specificity, predictive values and likelihood ratios. 2020.

26. World Health Organization, Female genital schistosomiasis: a pocket atlas for clinical health-care professionals. 2015, World Health Organization: Geneva, Switzerland.

27. Hendriks, M.E., et al., Step-by-step guideline for disease-specific costing studies in low- and middle-income countries: a mixed methodology. Glob Health Action, 2014. 7: p. 23573.

28. Turner, H.C., et al., What are economic costs and when should they be used in health economic studies? Cost Effectiveness and Resource Allocation, 2023. 21(1): p. 31.

29. Financial Times, FT Guide to World Currencies, Morningstar, Editor. 2026.

30. Lamberti, O., Cost-effectiveness of screening strategies for female genital schistosomiasis in endemic settings. 2025, London School of Hygiene & Tropical Medicine.

31. Darlington, M., et al., Decrementally cost-effective health technologies in non-inferiority studies: A systematic review. Front Pharmacol, 2022. 13: p. 1025326.

32. StataCorp, Stata Statistical Software: Release 16.1. 2020, StataCorp LLC: College Station, TX.

33. R. Core Team, R: A Language and Environment for Statistical Computing. 2025, R Foundation for Statistical Computing: Vienna, Austria.

34. Mbwanji, G., et al., High sensitivity but low specificity of the risk factors and symptoms questionnaire in diagnosing female genital schistosomiasis among sexually active women with genital lesions in selected villages of Maswa District, North-Western Tanzania. PLoS Negl Trop Dis, 2024. 18(8): p. e0012336.

35. Aribodor, O.B., et al., Assessing urogenital schistosomiasis and female genital schistosomiasis (FGS) among adolescents in Anaocha, Anambra State, Nigeria: implications for ongoing control efforts. BMC Public Health, 2024. 24(1): p. 952.

36. Wall, K.M., et al., Imported female genital schistosomiasis: a neglected health issue across borders. J Glob Health, 2026. 16: p. 03002.

37. Mbwanji, G., et al., Female genital schistosomiasis is a neglected public health problem in Tanzania: Evidence from a scoping review. PLOS Neglected Tropical Diseases, 2024. 18(3): p. e0011954.

38. Swai, B., et al., Female genital schistosomiasis as an evidence of a neglected cause for reproductive ill-health: a retrospective histopathological study from Tanzania. BMC Infect Dis, 2006. 6: p. 134.

39. Kjetland, E.F., et al., Female genital schistosomiasis – a differential diagnosis to sexually transmitted disease: genital itch and vaginal discharge as indicators of genital Schistosoma haematobium morbidity in a cross-sectional study in endemic rural Zimbabwe. Tropical Medicine & International Health, 2008. 13(12): p. 1509–1517.

40. Oluwole, A.S., et al., A quality improvement approach in co-developing a primary healthcare package for raising awareness and managing female genital schistosomiasis in Nigeria and Liberia. International Health, 2023. 15(Supplement_1): p. i30–i42.

41. Campos, N.G., et al., Cervical cancer screening in low-resource settings: A cost-effectiveness framework for valuing tradeoffs between test performance and program coverage. Int J Cancer, 2015. 137(9): p. 2208–19.

42. Webb, J.A., et al., The cost-effectiveness of schistosomiasis screening and treatment among recently resettled refugees to Canada: an economic evaluation. CMAJ Open, 2021. 9(1): p. E125–e133.

43. Banda, H. and A. Abere, Determining the mechanisms of praziquantel resistance in schistosomiasis: insights and future directions. Discover Medicine, 2025. 2(1): p. 105.

44. Arenholt, L.T.S., et al., Repeated versus single praziquantel dosing regimen in treatment of female genital schistosomiasis: a phase 2 randomised controlled trial showing no difference in efficacy. Frontiers in Tropical Diseases, 2024. Volume 5 - 2024.

